# Seasonality of Respiratory, Enteric and Urinary Viruses Revealed by Wastewater Genomic Surveillance

**DOI:** 10.1101/2024.02.06.24302386

**Authors:** Matthew F. Smith, Rabia Maqsood, Regan A. Sullins, Erin M. Driver, Rolf U. Halden, Efrem S. Lim

## Abstract

Wastewater surveillance can reveal population-level infectious disease burden and emergent public health threats can be reliably assessed through wastewater surveillance. While molecular methods for wastewater monitoring of microorganisms have traditionally relied on PCR-based approaches, next-generation sequencing can provide deeper insights via genomic analyses of multiple diverse pathogens. We conducted a year-long sequencing surveillance of 1,408 composite wastewater samples collected from 12 neighborhood-level access points in the Greater Tempe area, Arizona, USA, and show that variation in wastewater viromes is driven by seasonal time and location. Wastewater virome temporal dynamics were influenced in a cyclical manner, with the most dissimilarity between samples 23 weeks apart (i.e., winter vs summer, spring vs fall). We identified diverse urinary and enteric viruses including polyomaviruses, astroviruses and noroviruses, and showed that their genotypes/subtypes shifted across season. We show that while wastewater data of certain respiratory viruses like SARS-CoV-2 strongly correlate with clinical case rates, laboratory-reported case incidences were discordant with surges of high viral load in wastewater for other viruses like human coronavirus 229E. These results demonstrate the utility of wastewater sequencing for informing decision making in public health.

**IMPORTANCE:** Wastewater genomic sequencing surveillance can provide insights into the spread of pathogens in communities. Advances in next-generation sequencing methodologies allow for more precise detection of viruses in wastewater. Long-term wastewater genomic sequencing surveillance is an important tool for public health preparedness. This system can act as a public health observatory that gives real-time early warning for infectious disease outbreaks and improved response times.

## INTRODUCTION

Wastewater genomic surveillance can be an effective tool for public health monitoring of pathogens. Wastewater constitutes a complex chemical and biological matrix representative of contributing human, animal, and microbiological communities. Hence, wastewater monitoring of a catchment can track health threats in the catchment community in a non-invasive and cost-effective manner. Wastewater-based epidemiology has been used to analyze the presence of microbes (1, 2), antimicrobial resistance genes (3), and chemical biomarkers (4–6). These data can be used to better understand population-level health and assess emerging public health risks. During the COVID-19 pandemic, wastewater genomic surveillance was used to track severe acute respiratory syndrome coronavirus 2 (SARS-CoV-2) circulating in communities. Wastewater-based epidemiology was employed to monitor outbreaks and community transmission of SARS-CoV-2 (7, 8).

PCR-based analysis of wastewater for SARS-CoV-2 (e.g., qRT-PCR, RT-dPCR) can supplement public health data by providing real-time data on population-level disease prevalence and has been shown to be less prone to sampling bias when compared to clinical surveillance of individuals (9). Furthermore, genomic sequencing of wastewater provided information about circulating SARS-CoV-2 variants by revealing cryptic lineages previously unobserved in clinical surveillance (10, 11) and early warning of emerging SARS-CoV-2 variants of concern (12, 13). This showcased the potential for wastewater surveillance in monitoring emergent pathogen threats (14–16).

Wastewater-based epidemiology has been demonstrated for other viral pathogens such as influenza virus, respiratory syncytial virus (RSV) and human rhinovirus (17–19). While these wastewater genomic studies traditionally rely on targeted PCR-based readouts (17–19), high-throughput next-generation sequencing methodologies are uniquely suited to interrogate complex microbial communities in wastewater samples (20, 21). Given the complex wastewater matrix, hybrid-capture enrichment can allow for robust virus detection (22, 23), whereas shotgun metagenomic sequencing has very little success in capturing wastewater-borne viruses (24). Thus, use of next-generation sequencing and bioinformatics approaches promises to reveal more informative and multifaceted insights into pathogens (evolution, diversity, genomic mutations, etc.) beyond simply detection and quantification.

Seasonal patterns in respiratory and enteric pathogenic virus infections have been observed (25–32). In the United States, the National Respiratory and Enteric Virus Surveillance System (NRVESS) lab-based reporting shows that respiratory viruses such as coronaviruses (aside from SARS-CoV-2), RSV and human metapneumovirus, occur in late fall, winter and/or early spring. Other respiratory viruses such as human adenovirus generally do not exhibit seasonal pattern but are circulating throughout the year (28, 33). However, certain subtypes of human adenovirus that can cause conjunctivitis or gastrointestinal disease may exhibit seasonality (29, 30). Enteric viruses such as human norovirus also occur more frequently during late fall, winter, and early spring seasons (27, 31, 32). Environmental factors and human behavior are the main contributors to the seasonality of viruses (34–37). Here, in a year-long wastewater surveillance study, we show the occurrence of virus dynamics in a seasonal patterns which eluded conventional public health surveillance networks.

## RESULTS

### One-year wastewater sequencing surveillance of viruses in Tempe, Arizona

We initiated a city-level public health genomic sequencing “Arizona Health Observatory” surveillance system to track viruses in wastewater weekly over the period of one year across 2022. In this study, 1,408 wastewater samples were collected from 12 locations (**Figure 1A**) three times a week in the Greater Tempe region in Arizona (population size of approximately 700,000 people). The sampling locations represent catchments with population estimates of 505,000 to 6,500 residents (AZ01–AZ09; sampled from January 6^th^, 2022, through December 29^th^, 2022), and 3 locations (AZ10 and AZ11; sampled from January 6^th^, 2022, through September 10^th^ 2022. AZ12; sampled January 6^th^, 2022, through September 24^th^, 2022) representing small, transient use catchments for which their service area population could not be estimated with confidence. (**Supplementary Figure 1A**).

**Figure 1.**
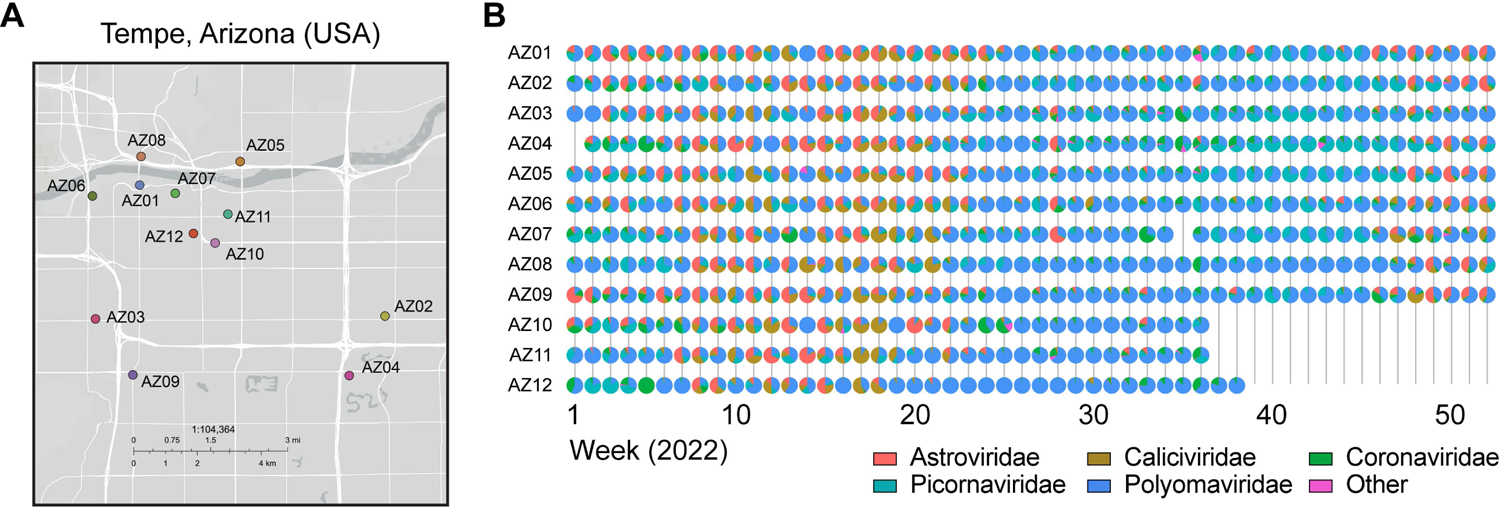
Wastewater genomic sequencing surveillance study design. (A) Map of Greater Tempe Area with locations of 12 sampling sites specified by colored circles. (B) Family-level relative abundance of reads per million normalized reads (rows: sampling location, columns: weeks).

Total nucleic acid (DNA and RNA) was extracted from samples and pooled by collection week for each location resulting in a total of 576 pooled samples. Median viral loads of pepper mild mottle virus (PMMoV), a common human fecal indicator virus, in pooled samples were within expected ranges for untreated wastewater (38) (Median concentration 3.44 x 10^7^ copies per liter wastewater; **Supplementary Figure 1B**). Libraries were prepared using hybrid capture enrichment for a panel of 66 DNA and RNA virus targets of high public health concern (Illumina Viral Surveillance Panel). Sequencing generated an average of 25.2 million paired reads per sample (average of 7.5 million QC-filtered reads per sample). Sequencing reads were quality-filtered and mapped to a custom database of virus genomes comprised of the enrichment panel targets, normalized per million reads. The most common viruses detected at the family taxonomic level were *Polyomaviridae* (52.7% average relative abundance), *Picornaviridae* (15.9%), *Astroviridae* (13.3%), *Caliciviridae* (10.8%) and *Coronaviridae* (6.3%) (**Figure 1B**).

### Influence of seasonality on wastewater virome diversity and distribution

Because respiratory viral infections have seasonal patterns (28, 39), we tested the hypothesis that seasonality influences wastewater virome diversity. Using linear mixed effects models, we found that virome alpha diversity (Shannon index) was significant altered by season and week (seasons *P* = 2.2 x 10^-16^, week *P* = 0.0043; **Figure 2A**), decreasing to the lowest Shannon diversity in the summer and increasing to the highest in the winters. We next compared the variability of viromes among wastewater samples by quantifying beta diversity measurements. Principal coordinates analysis (PCoA), as measured by Bray-Curtis distances, showed that wastewater samples differed primarily by season and secondly by week (PERMANOVA seasons *P* < 0.001, week *P* < 0.05, **Figure 2B**). The median Bray-Curtis dissimilarity was significantly lower within the same season than between seasons (*P* < 0.0001, **Supplementary Figure 1C**), and within samples from the same sampling location than between different collection sites when controlling for season (*P* < 0.0001, **Supplementary Figure 1D**), indicating that both time and location contribute to wastewater virome variation. To confirm the cyclical pattern in wastewater viromes (**Figure 2B**, Axis 1), we compared Bray-Curtis distances as a function of the week interval between samples. Virome dissimilarity in general was most pronounced when comparing samples that were collected 23 weeks apart (median dissimilarity 0.60, **Figure 2C, Supplementary Figure 1E**). Samples that were either closer in time or more than 40 weeks apart had higher similarity (i.e., lower Bray-Curtis distance). This suggests that temporal dynamics of wastewater viromes are influenced in a cyclical manner.

**Figure 2.**
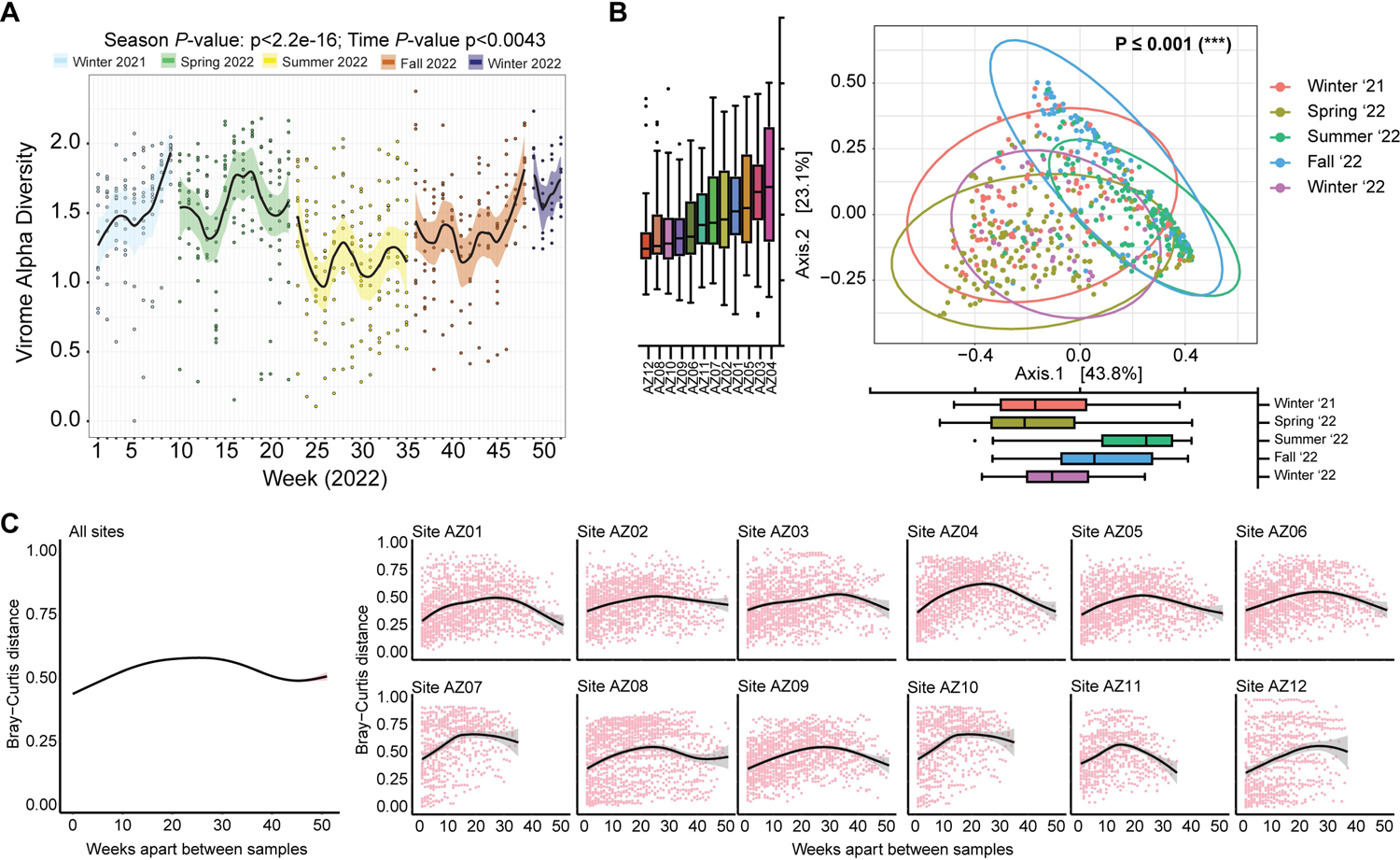
Seasonality of wastewater virome diversity. (A) Alpha diversity (Shannon Index) for each sample denoted by dots and linear mixed effects model with confidence interval shaded by meteorological season overlay. (B) Weighted beta diversity (bray-curtis) plotted as PCoA. Box plots of seasons (PC1) and sites (PC2). Color in PCoA represent the season and year. Statistical significance of variation in beta diversity explained by eteorological season assessed by a partial sums of squares PERMANOVA model with permutation constrained within strata of sampling site. (C) Weighted bray-curtis dissimilarity plotted by time difference in weeks between samples. Summary plot combined data for all sites across weeks apart, while subsequent plots are subset for individual sites across weeks apart.

To define the wastewater virome community structure, we applied k-means clustering and identified 4 community state types (**Figure 3**). Cluster 1 was most abundant in Astrovirus (37%), JC Polyomavirus (19%) and Aichivirus A (10%) and was associated with samples collected in spring (51%) and winter of 2021 (32%) (**Supplementary Figure 1F**). The second cluster was dominant in JC Polyomavirus (75%) and had 56% samples from the summer season (**Supplementary Figure 1E**). The third cluster that was abundant in JC Polyomavirus (39%), BK Polyomavirus (19%) and Aichivirus A (16%), and fourth cluster abundant in Aichivirus A (36%), JC Polyomavirus (18%) and SARS-CoV-2 (13%) were not significantly associated with seasons. In conclusion, alterations in wastewater viromes were associated with both the meteorological seasonality and sampling location.

**Figure 3.**
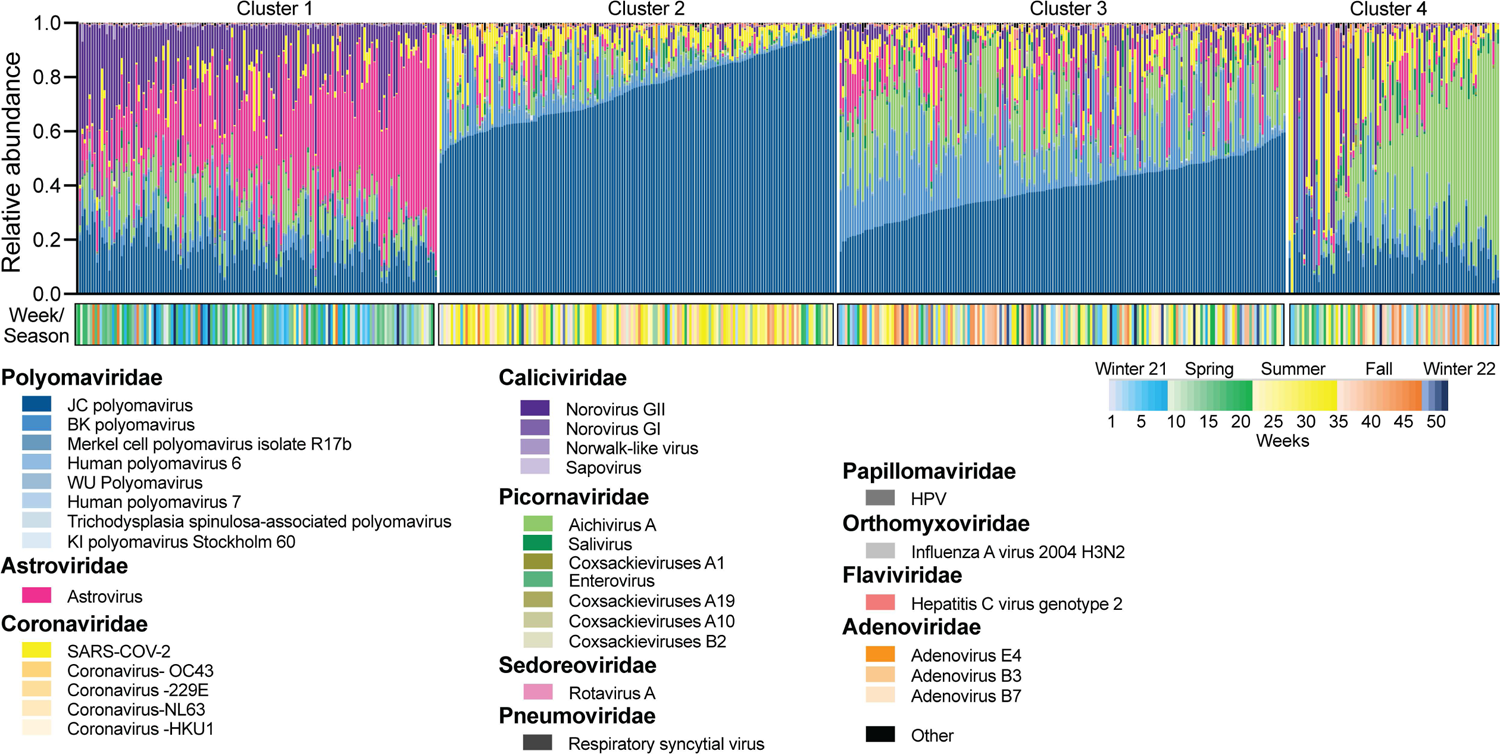
Community states and relative abundance of wastewater viruses. (A) Relative abundance of viruses at species level, clustered using k-means on weighted bray-curtis distances. Plot labeled with community state clusters. Clusters ordered by most abundant virus present Color bar at bottom of relative abundance plot represents 52 weeks (gradient) and seasons (color).

### Tracking respiratory viruses in wastewater

Wastewater levels of SARS-CoV-2 tracked with community disease prevalence during the COVID-19 pandemic (40–42). During our year-long 2022 surveillance study, SARS-CoV-2 case rates in Tempe strongly correlated with wastewater viral load (**Figure 4A** and **4B**, linear regression with Kendall’s rank correlation τ = 0.593). We compared the distribution of SARS-CoV-2 variants inferred from wastewater sequencing data to our ongoing baseline genomic surveillance efforts (13) in Tempe communities (n = 32,891 sequenced cases). The three major waves of Omicron BA.1, BA.2 and BA.5 in the population was correspondingly observed in wastewater lineage calls (**Figure 4C**). Additional minor lineage groups were observed in wastewater. Thus, the wastewater data reflected contemporaneous SARS-CoV-2 variants circulating in Tempe communities.

**Figure 4.**
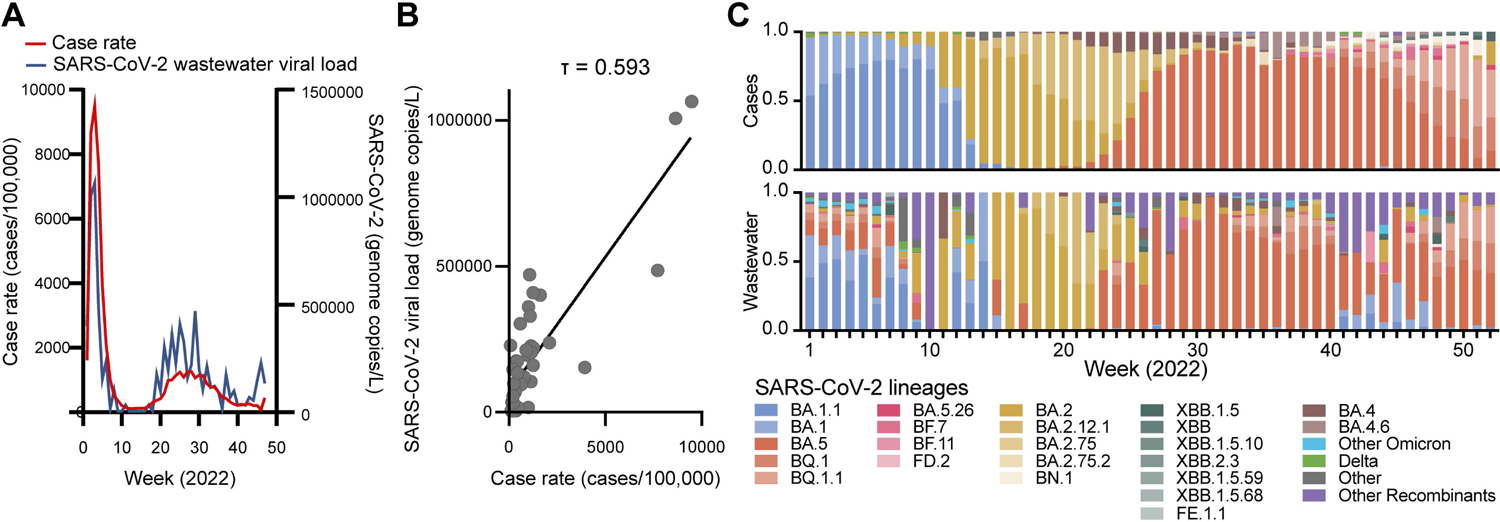
SARS-CoV-2 variant and viral load surveillance. (A) SARS-CoV-2 case rate (cases per 100,000) in Tempe (left) and median viral load (genome copies/L) measured from 6 publicly reported sites included in this study (right). (B) Linear regression and Kendall’s ranked correlation T (tau) between SARS-CoV-2 case rate (cases per 100,000) in Tempe (x-axis and median viral load (genome copies/L). (C) Weekly relative abundance of parental lineage groups from Arizona SARS-CoV-2 genomic surveillance (top) and weekly average relative abundance of lineage groups aggregated from Freyja lineage proportions.

In addition to SARS-CoV-2, other respiratory viruses detected by wastewater sequencing included human coronaviruses types 229E, NL63, OC43 and HKU1, human adenovirus, influenza A virus, human rhinovirus, human parainfluenza virus, respiratory syncytial virus, human parechovirus, and human metapneumovirus (**Figure 5A**). Typically, a common human alphacoronavirus and betacoronavirus is dominant each year (43). Hence, we compared wastewater data to clinical surveillance data for region 4 (west) from the National Respiratory and Enteric Virus Surveillance System (NREVSS) (**Figure 5B** and **5C**). Betacoronavirus HCoV-OC43 wastewater viral load was higher than HCoV-HKU1 in a manner consistent with NREVSS case data. Although alphacoronavirus HCoV-229E and HCoV-NL63 NREVSS cases rates were similar, levels of HCoV-229E in wastewater were higher than those of HCoV-NL63 suggesting a potential underreporting of HCoV-229E cases.

**Figure 5.**
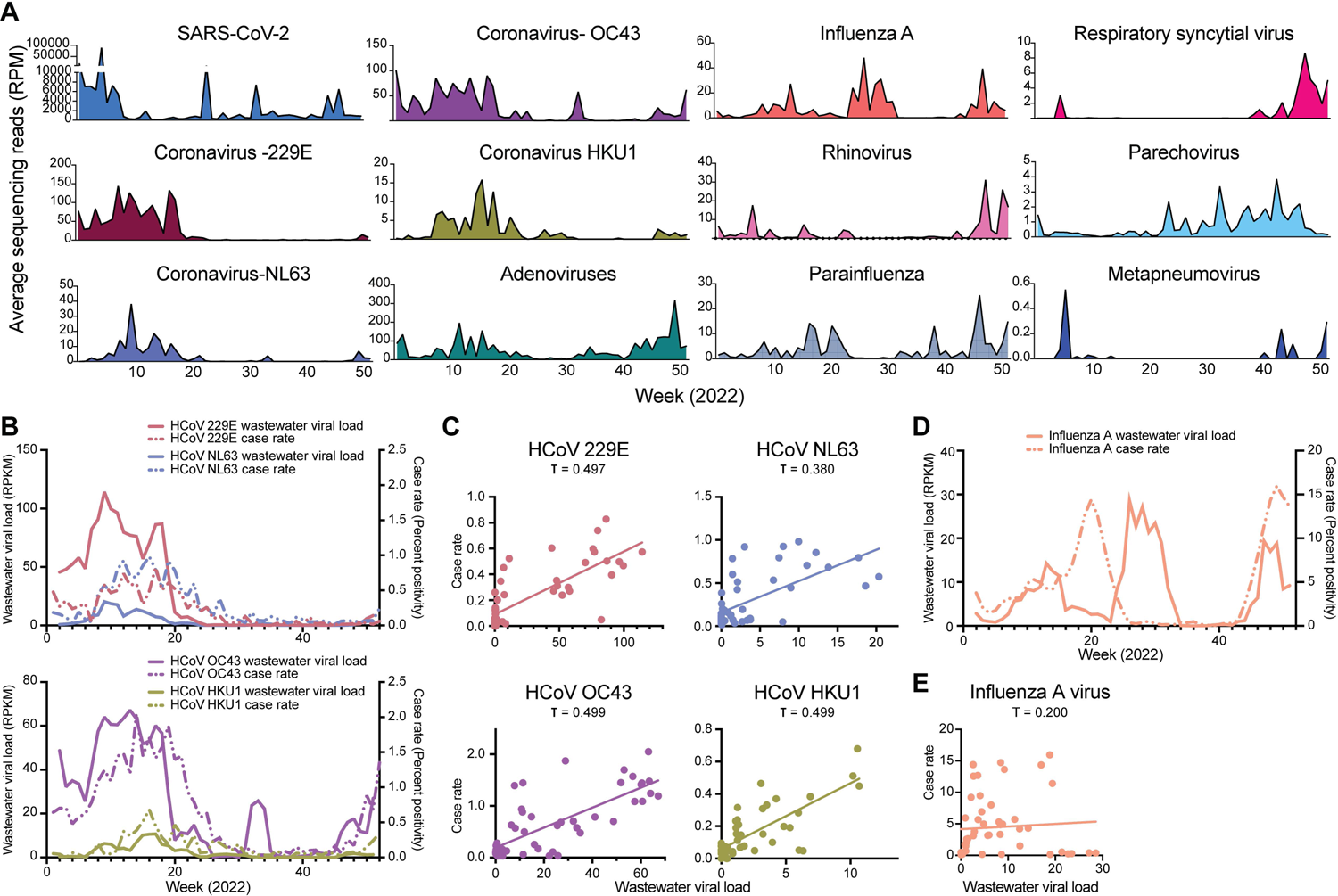
Respiratory viruses in wastewater. (A) Average sequencing reads of respiratory viruses found in wastewater plotted over 52 weeks. (B) 3-week centered average of wastewater normalized reads per million aligned to each seasonal coronavirus (left y-axis) and NREVSS Census Region West 3-week centered average case rate (testing percent positive) for each seasonal coronavirus (right y-axis) (top: Alphacoronavirus, bottom: Betacoronavirus). (C) Linear regression and Kendall’s ranked correlation **T** (tau) between 3-week centered average of wastewater normalized reads per million aligned to each seasonal coronavirus and NREVSS Census Region West 3-week centered average case rate (testing percent positive) for the same viruses. (D) 3-week centered average of wastewater normalized reads per million aligned to Influenza A (left y-axis) and CDC FluView testing case rate (percent positive) for Arizona. (E) Linear regression and Kendall’s ranked correlation T (tau) between 3-week centered average of wastewater normalized reads per million aligned to Influenza A and CDC FluView testing case rate (percent positive) for Arizona.

PCR-based detection of influenza A virus in wastewater has been previously observed to be associated with case incidence (44–46). Although 3 surges of influenza A virus were detected in wastewater in this study, the peak of wastewater detection between weeks 26 and 31 was offset when compared to state-level influenza A virus positivity rates in Arizona (**Figure 5D**). Linear regression and Kenall’s rank correlation analysis corroborated that the load of influenza A in wastewater correlated poorly with clinical positivity rate (**Figure 5E**; τ = 0.200). Overall, while certain infectious diseases are being tracked effectively, discordant findings may indicate gaps in surveillance.

### Differential distribution of polyomaviruses in wastewater

Polyomaviruses, DNA viruses associated with diseases such as BK polyomavirus nephropathy, progressive multifocal leukoencephalopathy and Merkel cell carcinoma, are ubiquitous in populations and can be shed in various ways including via urine, skin and stool (47). Hence, we investigated the wastewater-based epidemiology of polyomaviruses as a potential surveillance indicator. When compared to detection of SARS-CoV-2 (median reads per million = 457), JC polyomavirus (JCPyV) and BK polyomavirus (BKPyV) were detected at relatively high levels (median reads per million JCPyV = 8314, BKPyV = 882) (**Figure 6A**). Merkel cell polyomavirus, KI polyomavirus, WU polyomavirus, Trichodysplasia spinulosa associated polyomavirus, human polyomaviruses 6, 7 and 9 were detected at relatively intermediate or low levels throughout the year. BK polyomavirus subtypes are distributed geographically, with subtypes I and IV predominantly circulating in the United States (48). We deconvoluted the BKPyV subtypes by mapping sequencing reads to the VP1 gene and found that BKPyV subtype I was the dominant genotype, composed of subtypes Ia (average 73.8%), Ib-1 (13.2%), Ib-2 (9.8%), and Ic (0.5%) (**Figure 6B**). Unexpectedly, BKPyV subtypes II and III thought to be rarely found in the US were detected sporadically at low abundances (1.6% and 1.1% respectively), and subtype IV was rarely detected (IVa-1 and IVc-2; <0.01%). These results demonstrate that wastewater sequencing surveillance can reveal trends in the geographical distribution of polyomavirus genotypes within populations.

**Figure 6.**
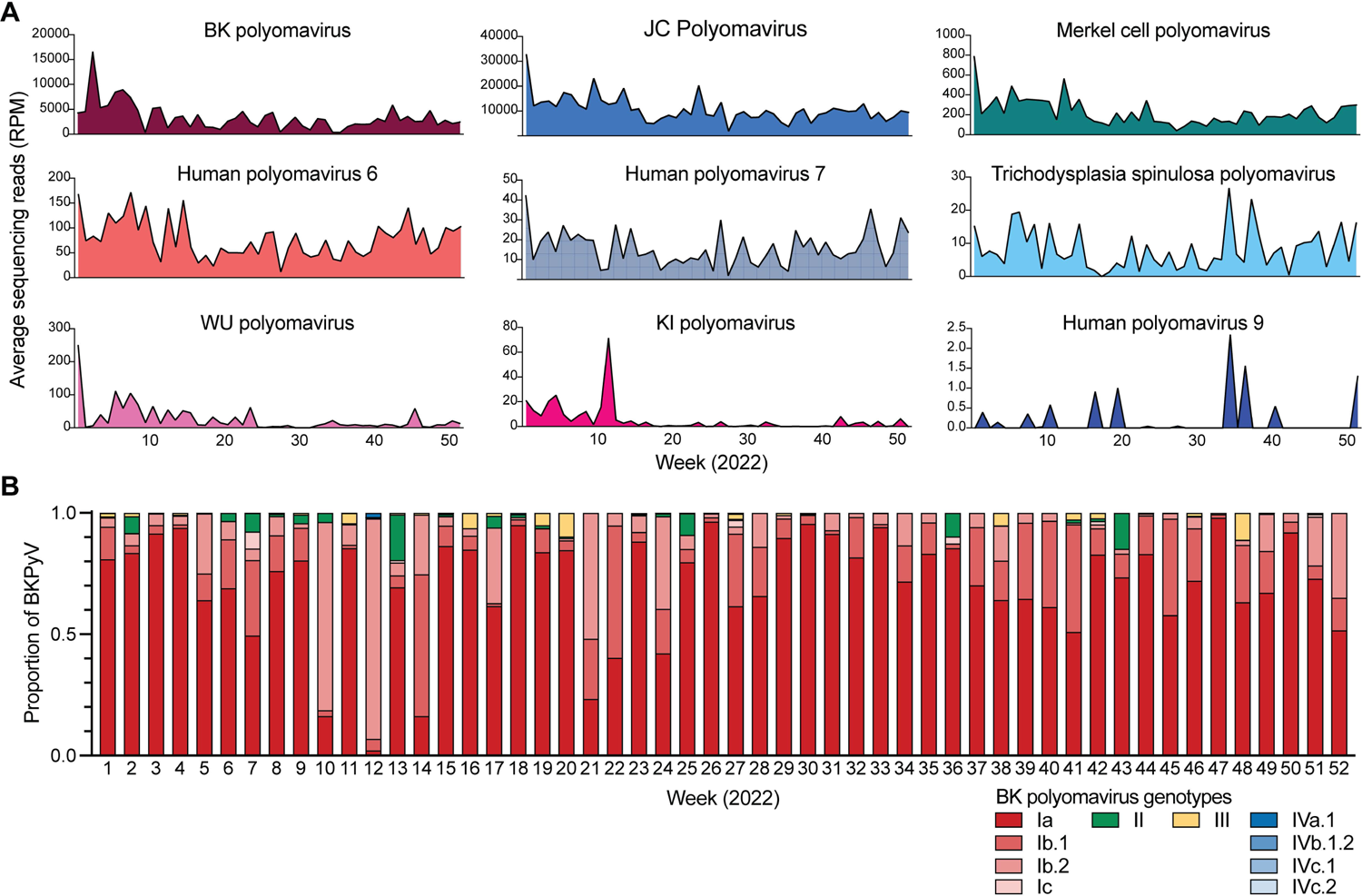
Polyomaviruses in wastewater. (A) Average sequencing reads of polyomaviruses found in wastewater plotted over 52 weeks. (B) BK Polyomavirus genotypes relative abundance plotted over 52 week study period.

### Monitoring human enteric viruses

We identified a diverse range of human enteric viruses in wastewater such as astrovirus, norovirus, aichivirus, salivirus, coxsackievirus, enterovirus, sapovirus and rotavirus (**Figure 7A**). Besides human astroviruses, a leading cause of acute gastroenteritis, mammalian and avian species also harbor diverse astroviruses that are generally host species-specific (49). By querying sequencing reads against a custom database of astrovirus capsid sequences, we predominantly detected human astroviruses (e.g., mamastrovirus 1, human astrovirus 1-8) (**Figure 7B**). Additionally, mammalian (cat and dog) and avian (bird) astroviruses were consistently detected in wastewater indicating that the wastewater signals included animal sources in the catchment. Tracking population-level genotypes of norovirus, another common cause of acute gastroenteritis, has important implications for norovirus vaccines in development (50). Hence, we developed a norovirus bioinformatics workflow for wastewater sequencing data and show that Norovirus GII and Norovirus GII.2 were the most abundant genotypes (**Figure 7C**). Further, we observed the proportion Norovirus GII.2 increased in late winter and spring. Interestingly, wild boar norovirus was detected, particularly during the summer and fall, suggesting the potential for human-swine interactions. Taken together, these findings demonstrate that wastewater sequencing can provide genomic resolution insights into emerging pathogen threats to better inform public health protection.

**Figure 7.**
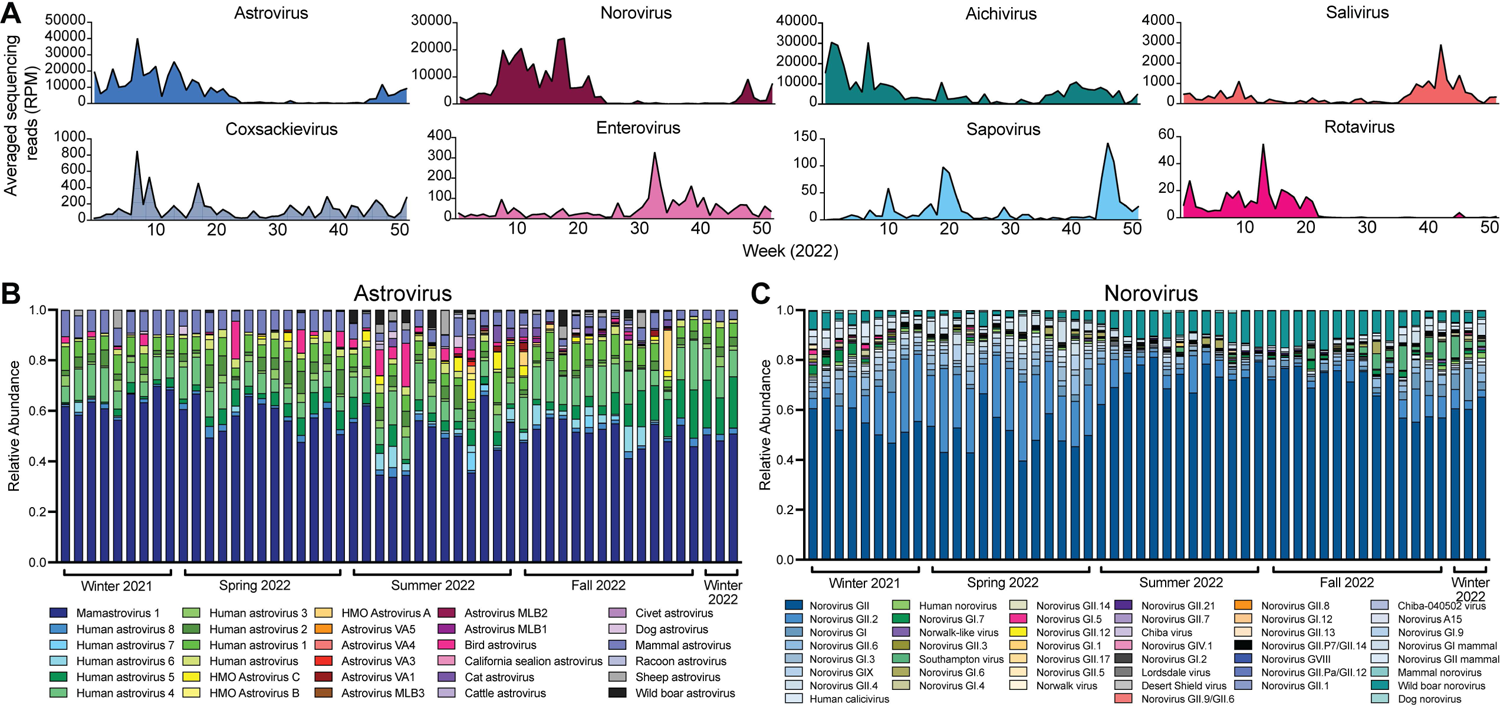
Enteric viruses in wastewater. (A) Average sequencing reads of enteric viruses found in wastewater plotted over 52 weeks. (B) Enteric pathogen Astrovirus abundance over the 52-week period. Presence of non-human Astrovirus found in wastewater aggregated by viral host species. (C) Enteric pathogen Norovirus abundance over the 52-week period. Presence of non-human Norovirus found in wastewater aggregated by viral host species.

## DISCUSSION

Wastewater surveillance can enhance monitoring of public health threats in populations. We demonstrate how deeper insights gleaned from next-generation sequencing applied in the context of urban wastewater surveillance can better inform public health decision making, compared to traditional PCR-based surveillance. The ability to interrogate diverse DNA and RNA viral targets in massively parallel wastewater sequencing represents a significant improvement that can contribute to pandemic preparedness and One Health approaches to tackling global health challenges (15). The advantages of wastewater surveillance include the unbiased and non-invasive nature of this approach that can be applied in communities where clinical surveillance data is sparse, and there are undiagnosed asymptomatic or mild disease transmissions. Next-generation sequencing approaches can be pathogen agnostic, making wastewater surveillance especially relevant to pandemic preparedness for novel and emerging pathogens and yet-to-be-characterized Pathogen X (51). The scope of surveillance can be tailored according to other strategic interests, such as human-animal interface of potential zoonotic transmissions, animal husbandry and agricultural sites. Overall, this wastewater surveillance framework relying on genomic sequencing was shown to generate rich, high-dimensional data that opens new avenues to study burden of infectious diseases with reduced bias at the community and population levels and to better understand factors that influence these patterns (e.g., human mobility, meteorological conditions).

Wastewater virus load and genomic variant/genotype distributions can correlate well with clinical disease incidence. In our study and others, SARS-CoV-2 shows robust correlation between wastewater viral load and local per capita case rates (52). As testing behavior trends change, genomic sequences of circulating SARS-CoV-2 variants can still be identified through wastewater sequencing to monitor important evolutionary dynamics. These genomic insights can inform public health and clinical practice decisions such as when new variants emerge harboring mutations that render monoclonal antibody therapeutics ineffectual (53, 54). For certain viral pathogens, our sequencing results are discordant with case prevalence. There are two possible explanations: first, as case prevalence was based on Arizona state-level data, this may be due to the limited catchment surveillance of Tempe city. Nonetheless, this emphasizes the importance of local public health responses. Second, as laboratory testing is not routinely performed for viruses such as common human coronaviruses (e.g., HCoV-229E), this could lead to underreporting and thus appear to be poorly correlated with wastewater signals. In fact, most viral infectious diseases are not tested as extensively as SARS-CoV-2 during the COVID-19 pandemic. These findings suggest that wastewater surveillance can reveal vulnerabilities in public health surveillance systems, even retrospectively.

There are several implications to our findings that high levels of BK polyomavirus and JC polyomavirus are detected in wastewater throughout the year. Since both BK polyomavirus and JC polyomavirus are human host-specific (55), commonly acquired during childhood (56), shed in urine (57), and ubiquitous in populations including healthy, immunocompetent and immunocompromised individuals (58, 59), they could be alternative biomarkers for data normalization in wastewater-based surveillance.

While the predominance of BK polyomavirus subtype I (Ia, Ib-1, Ib-2 and Ic) is consistent with its known geographic distribution in North America, subtypes II and III are rarely detected, making our findings unexpected. As BK polyomavirus is implicated in nephropathy and graft loss, most studies have been focused on renal transplantation, and genotyping particularly of transplant recipients and donors. This suggests that a better understanding of BK polyomavirus diversity and population distributions can be achieved through wastewater surveillance approaches. A caveat of wastewater next-generation sequencing is that the sequencing data is the genomic aggregate of large numbers of individuals, making it inappropriate and/or difficult to accurately assemble individual virus genome sequences. However, this challenge could be overcome with advancements in long-read sequencing technologies.

Our long-term wastewater surveillance in th Greater Tempe area, Arizona, shows the seasonal patterns of human viral pathogens over the year. Seasonal trends have also been observed of viruses in other wastewater studies (23, 31, 60). Due to the shorter study period that did not encapsulate a full year, the cyclical behavior of the wastewater virome was only partially observed in a previous study (23). Our results indicate that wastewater viromes are influenced in a cyclical manner. We show that wastewater virome diversity was most dissimilar when comparing to other samples collected 23 weeks apart (e.g., spring vs fall, winter vs summer), whereas samples collected 40-51 weeks apart cycling back to the same season were more alike (i.e., low beta diversity). While these results show that the viral community in winter 2021 resembles winter 2022, future multi-year studies are needed to better define the regularity of the cyclical behavior in wastewater viromes. Our findings advance our understanding of the seasonal patterns of viruses and demonstrate the importance of wastewater surveillance in public health. Throughout the 52 weeks of this study, we were effectively able to find high diversity of viruses specific to enteric, urinary, and respiratory regions. There were both commensal and pathogenic viruses in the wastewater and we also detected specific trends for association between seasons and viruses present.

## MATERIALS AND METHODS

### Sample collection and processing

A total of 1,408 composite samples of untreated wastewater were collected over 24 hours from within the wastewater collection system using high frequency automated samplers at 12 locations in the Greater Tempe area, Arizona, USA. Samples were primarily collected on Tuesdays, Thursdays, and Saturdays (three times weekly). Nine sampling locations (AZ01–AZ09) were sampled for the full year period (January 6^th^, 2022 through December 29^th^, 2022) generating between 119-131 distinct wastewater samples per location. Three sampling locations (AZ10–AZ12) were sampled for approximately 70% of the year (AZ10 and AZ11: January 6^th^, 2022, through September 10^th^ 2022. AZ12: January 6^th^, 2022, through September 24^th^, 2022) generating between 87-89 distinct wastewater samples per location.

Samples were transferred to 1L high-density polyethylene (HDPE) bottles and stored at 4°C until concentration and nucleic acid extraction. Wastewater samples were processed as previously described (10). Briefly, raw wastewater was filtered through a 0.45 µm polyethersulfone (PES) membrane filter (Fisher Scientific, Lenexa, KS, USA). Viruses present in the resultant solids-depleted filtrate were concentrated on an Amicon ultra 15 centrifugal filter with a 10,000 molecular weight cutoff (MWCO) (Millipore Sigma, Burlington, MA, USA). DNA and RNA was extracted from the concentrate using a Qiagen RNeasy Mini Kit (Qiagen, Germantown, MD, USA). Nucleic acids were pooled according to sampling location and week of collection date (weeks beginning Sunday and ending Saturday following the MMWR week numbering convention; week 1 starting 1/2/2022 and week 52 ending 12/31/2022). Pooling resulted in a total of 576 pooled samples—51-52 pools for the sites AZ01–AZ09 sampled for the full year and 36-38 pools for the sites AZ10–AZ12 sampled for the partial year.

### Next generation sequencing (NGS)

NGS library preparation was performed using the Illumina RNA Prep with Enrichment kit Viral Surveillance Panel (Illumina, San Diego, CA, USA). Libraries were sequenced on the Illumina NextSeq2000 instrument using 2 × 151 paired end reads generating an average of 25,281,226 paired end sequencing reads per sample and an average of 7,524,664 post-QC filtered paired end sequencing reads.

Demultiplexing of fastq files was performed with Illumina DRAGEN software BCL Convert (version 3.8.4). Fastq quality control workflow was performed with the BBTools suite (Bushnell 2016). Briefly, adapter trimming, read quality and length filtering, and contaminant removal (PhiX) were performed using BBDuk specifying a minimum length of 75 bases and minimum Phred Quality score of 20. Reads with at least 99% identity to another read were filtered out using Dedupe. Overlapping reads were joined using BBMerge before a second round of deduplication with Dedupe on merged reads specifying a minimum identity of 100% to demark duplicates. All remaining reads (deduplicated merged and unmerged read pairs) were passed through BBDuk a final time to remove BBDuk-default Illumina sequences.

Quality-filtered fastq files were converted to fasta format and reads were mapped to a custom database containing representative reference sequences for viruses included in the enrichment panel using Bowtie 2 (61) to generate SAM format alignments. Non-primary alignments, supplementary alignments, and unmapped reads were filtered from the primary alignment using SAMtools view (62). Primary SAM format alignments were converted to BAM format and indexed using SAMtools sort and index. Mapping statistics were computed from each primary BAM file using SAMtools idxstats. Calculation of normalized read counts (Reads Per Million; RPM) for all samples was performed in R statistical software (R Core Team 2020).

### Virome sequencing analysis

To determine the counts of each enteric, respiratory, and polyomavirus over the weeks, we plotted the averaged normalized counts across site by time. For Polyomaviruses, we were interested in characterizing the relative abundance of BK polyomavirus subgroups and subtypes in wastewater over the year. We applied a previously described algorithm adapted from the program BKTyper (63) to detect SNPs associated with each BK polyomavirus subgroup from only BK polyomavirus aligned reads which span the entire typing region within the VP1 gene. For enteric viruses, we also wanted to illustrate the unique species and variants of the two most abundant enteric pathogens in the wastewater. To do so, we created a custom database of the capsid protein region by downloading capsid protein regions for Norovirus and Astrovirus on NCBI Virus and mapped the QC reads with BWA to get counts of variant/species per sample. If multiple accession for same species/variants, we aggregated the counts by species name. If non-human viruses were found, we aggregated the counts by host species. We then normalized the counts to 250K (minimum qc read depth), from which we obtained relative abundance to plot across sites for each week.

### Pepper mild mottle virus (PMMoV) assay

We applied a previously described RT-qPCR assay targeting Pepper Mild Mottle Virus (PMMoV) (64) as a control to assess potential inhibition by residual wastewater contaminants and whether viruses shed in feces by healthy adults would be detectable across all wastewater samples. Thermal cycling conditions were performed as described in a QuantStudio 7 Flex Real-Time PCR System (Applied Biosystems, Waltham, MA, USA). Reactions were performed in a total volume of 25 µL consisting of 12.5 µL SuperScript III One-Step 2X Reaction Mix, 0.5 µL SuperScript III RT/Platinum Taq mix (Invitrogen, Waltham, MA, USA), 1 µL of forward primer PMMV-FP1-rev at a concentration of 10 µM, 1 µL of reverse primer PMMV-RP1 at a concentration of 10 µM, 0.5 µL probe PMMV-Probe1 at a concentration of 10uM, 4.5 µL nuclease-free water, and 5 µL template. Ten-fold serial dilutions of a 125bp synthetic DNA fragment spanning the target sequence were utilized to generate a standard curve to quantify PMMoV genome copies present in extracted samples. As DNA standards were used in place of RNA standards, reverse transcription efficiency was not considered in this assay and resulting PMMoV concentrations are potentially underestimated with the true concentration of PMMoV in wastewater samples likely to be higher.

### Linear mixed effects and PERMANOVA analysis

We used the R package lmerTest to create linear models using seasons and weeks as fixed effects variables and sites as random variables to control for multiple samples from each site. Using the models, we applied ANOVA and tested for changes in the alpha diversity and richness by seasons and time. Using the adonis2() function from R package vegan, we applied a partial sum of squares PERMANOVA model to the bray-curtis dissimilarity matrix to determine the variation in beta diversity associated with explanatory variables week and season while constraining permutation strata within sampling site.

To compare beta diversity across sites we controlled and used the weighted Bray-Curtis distance matrix to plot the distances by same seasons and same site and same seasons and different site and performed a Mann-Whitney test to test for significant differences between same site and different sites. We also wanted to check for differences across seasons and plotted the distances during the same season and between seasons across all sites and used Mann-Whitney to test for significant differences by seasons. To determine if there is cyclical relationship in the samples, we plotted the weighted beta diversity by the absolute differences in weeks between samples. If there were a cyclical relationship, we should see the sample with the least and the most weeks apart to be more similar than the samples midway between.

### K-means clustering and community state analysis

To obtain the community states present in the wastewater samples of this study we clustered our weighted beta diversity distance matrix with k-means method for the viruses. We first used the R package factoextra (version 1.0.7) to determine the optimal number of clusters and then used stats function k-means to cluster the samples into four groups. We plotted the species relative abundance level for each group and ordered the samples by the most abundant species present in that cluster. To determine associations between community states and seasons and time, we used R package mclogit (version 0.8.7.2) to perform multinomial logit models with random effects for sites; the Benjamini-Hochberg method was used to correct for multiple comparisons for the mclogit results.

### SARS-CoV-2 analysis

SARS-CoV-2 case rates were downloaded from a publicly available data dashboard reporting case rate for Tempe zip codes from the Maricopa County Department of Public Health. SARS-CoV-2 wastewater viral load was measured via RT-qPCR as previously described (40) and resulting data was obtained from the Tempe COVID-19 Wastewater Collection Data Dashboard. Aggregated SARS-CoV-2 genome copies per liter was calculated as the median genome copies per liter of wastewater across all sites within collection week. Individual-level SARS-CoV-2 genome sequences were generated from our ongoing baseline genomic sequencing surveillance efforts (13). A total of 32,891 viral genomes in 2022 were used for this analysis, all of which are made publicly available in the GISAID database. For wastewater samples, we analyzed relative lineage abundances using Freyja (12) and applied a genome breadth-of-coverage threshold of 50% for analysis. Of 576 pooled wastewater samples, 308 met the criteria for lineage analysis. Weekly lineage abundance was aggregated to major lineage groups.

### Respiratory virus analysis

The common coronaviruses 229E, NL63, OC43 and HKU1 and influenza A virus in wastewater were compared to clinical surveillance data reported by the National Respiratory and Enteric Virus Surveillance System (NREVSS) for Census Region 4 (West). The 3-week centered average of normalized reads mapped to each common coronavirus and the 3-week centered average NREVSS Census Region 4 positivity rate for each common coronavirus were analyzed.

## ABBREVIATIONS

SARS-CoV-2: Severe acute respiratory syndrome coronavirus 2

COVID-19: Coronavirus Disease 2019

qRT-PCR: Quantitative reverse transcription polymerase chain reaction

RT-dPCR: Reverse transcription digital polymerase chain reation

RSV: Respiratory syncytial virus

NRVESS: National Respiratory and Enteric Virus Surveillance System

PCoA: Principal coordinates analysis

DNA: Deoxyribonucleic acid

RNA: Ribonucleic acid

PMMoV: Pepper mild mottle virus

JCPyV: JC polyomavirus

BKPyV: BK polyomavirus

HDPE: High-density polyethylene

PES: Polyethersulfone

MWCO: Molecular weight cutoff

NGS: Next generation sequencing

RPM: Reads per million

## AVAILABILITY OF DATA AND MATERIALS

Sequencing data have been deposited to the NCBI Sequence Read Archive under accession number PRJNA1070654. Code used for diversity and ecological analyses is available at https://github.com/ASU-Lim-Lab/Wastewater-Genomic-Surveillance-Manuscript.

## AUTHOR CONTRIBUTIONS

Conceptualization: E.S.L.; Data curation: M.F.S., R.M.; Formal Analysis: M.F.S., R.M.; Funding acquisition: E.S.L.; Investigation: M.F.S., R.M., R.A.S., E.M.D.; Resources: E.M.D., R.U.H.; Supervision: E.S.L.; Visualization: M.F.S., R.M., E.S.L.; Writing – original draft: M.F.S., R.M., E.S.L. All authors reviewed and approved the final manuscript.

## Data Availability

All data produced in the present study are available upon reasonable request to the authors.

---

**Supplementary Figure 1.**
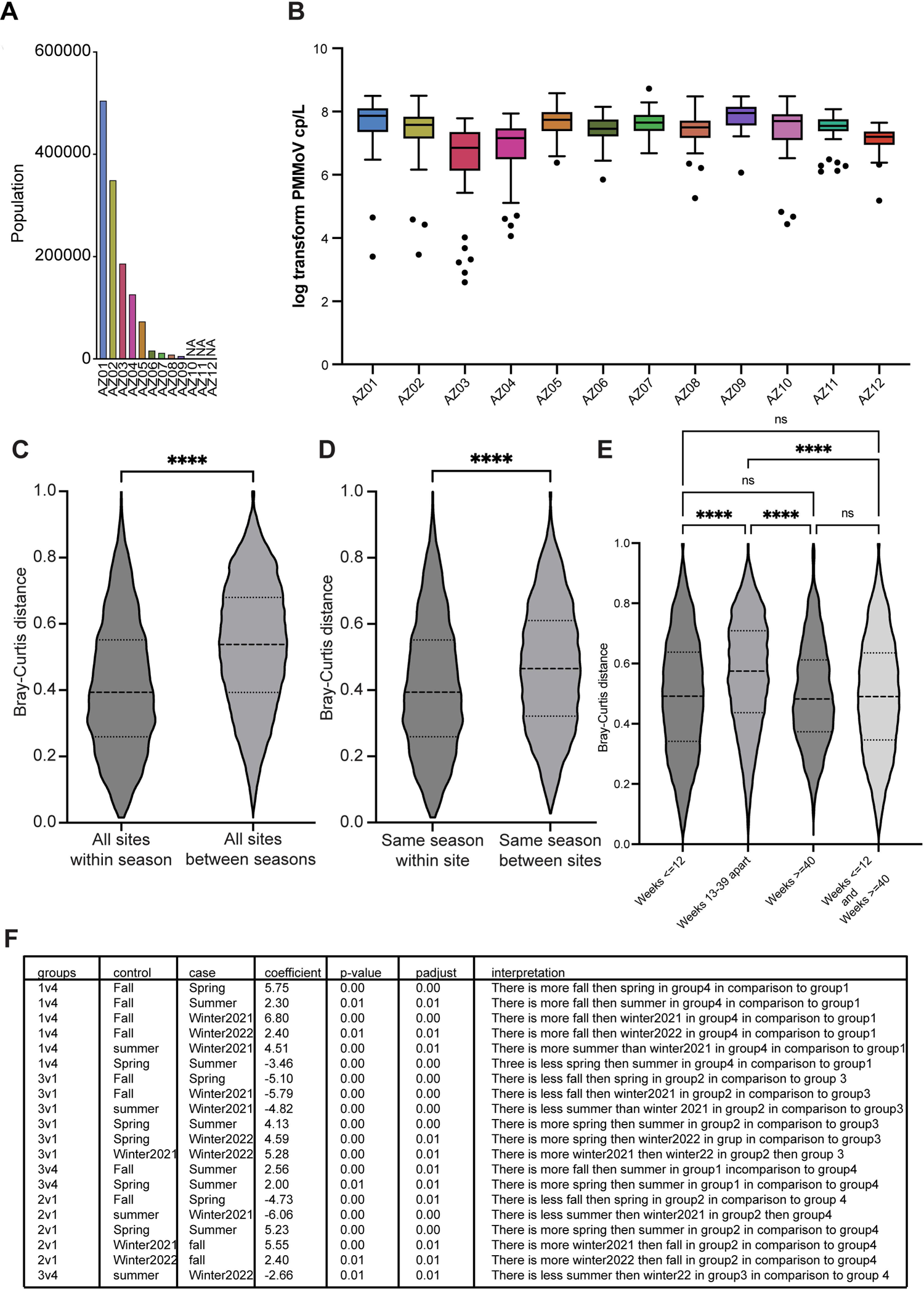
(A) Population estimates for catchments represented by each sampling site. (B) Box-plots of PMMoV viral load in wastewater (genome copies per liter wastewater) for each site. (C) Violin plot of bray-curtis distances of all sites by same season vs. different seasons. Statistical significance assessed by Mann-Whitney. (D) Violin plot of bray-curtis distances of same sites vs. different sites, controlled by same season. Statistical significance assessed by Mann-Whitney. **(E)** Bray-curits distances plotted for samples 1-12 weeks apart, 13-39 weeks apart, more than 40 weeks apart and combining first and third violin plot values. Statisti­ cal significance assessed by Kruskal Wallis with corrections for multiple comparisons. **(F)** Table for significant comparisons done using mclogit results for k-means clusters and metrological seasons.

## References

1. Gulino K, Rahman J, Badri M, Morton J, Bonneau R, Ghedin E. 2020. Initial Mapping of the New York City Wastewater Virome. mSystems 5.

2. LaMartina EL, Mohaimani AA, Newton RJ. 2021. Urban wastewater bacterial communities assemble into seasonal steady states. Microbiome 9:116.

3. Raven KE, Ludden C, Gouliouris T, Blane B, Naydenova P, Brown NM, Parkhill J, Peacock SJ. 2019. Genomic surveillance of Escherichia coli in municipal wastewater treatment plants as an indicator of clinically relevant pathogens and their resistance genes. Microb Genom 5.

4. Gushgari AJ, Venkatesan AK, Chen J, Steele JC, Halden RU. 2019. Long-term tracking of opioid consumption in two United States cities using wastewater-based epidemiology approach. Water Res 161:171–180.

5. Andres-Costa MJ, Proctor K, Sabatini MT, Gee AP, Lewis SE, Pico Y, Kasprzyk-Hordern B. 2017. Enantioselective transformation of fluoxetine in water and its ecotoxicological relevance. Sci Rep 7:15777.

6. Driver EM, Bowes DA, Halden RU, Conroy-Ben O. 2022. Implementing wastewater monitoring on American Indian reservations to assess community health indicators. Sci Total Environ 823:153882.

7. Lodder W, de Roda Husman AM. 2020. SARS-CoV-2 in wastewater: potential health risk, but also data source. Lancet Gastroenterol Hepatol 5:533–534.

8. Ahmed W, Angel N, Edson J, Bibby K, Bivins A, O’Brien JW, Choi PM, Kitajima M, Simpson SL, Li J, Tscharke B, Verhagen R, Smith WJM, Zaugg J, Dierens L, Hugenholtz P, Thomas KV, Mueller JF. 2020. First confirmed detection of SARS-CoV-2 in untreated wastewater in Australia: A proof of concept for the wastewater surveillance of COVID-19 in the community. Sci Total Environ 728:138764.

9. Nemudryi A, Nemudraia A, Wiegand T, Surya K, Buyukyoruk M, Cicha C, Vanderwood KK, Wilkinson R, Wiedenheft B. 2020. Temporal Detection and Phylogenetic Assessment of SARS-CoV-2 in Municipal Wastewater. Cell Rep Med 1:100098.

10. Fontenele RS, Kraberger S, Hadfield J, Driver EM, Bowes D, Holland LA, Faleye TOC, Adhikari S, Kumar R, Inchausti R, Holmes WK, Deitrick S, Brown P, Duty D, Smith T, Bhatnagar A, Yeager RA, 2nd, Holm RH, von Reitzenstein NH, Wheeler E, Dixon K, Constantine T, Wilson MA, Lim ES, Jiang X, Halden RU, Scotch M, Varsani A. 2021. High-throughput sequencing of SARS-CoV-2 in wastewater provides insights into circulating variants. Water Res 205:117710.

11. Smyth DS, Trujillo M, Gregory DA, Cheung K, Gao A, Graham M, Guan Y, Guldenpfennig C, Hoxie I, Kannoly S, Kubota N, Lyddon TD, Markman M, Rushford C, San KM, Sompanya G, Spagnolo F, Suarez R, Teixeiro E, Daniels M, Johnson MC, Dennehy JJ. 2022. Tracking cryptic SARS-CoV-2 lineages detected in NYC wastewater. Nat Commun 13:635.

12. Karthikeyan S, Levy JI, De Hoff P, Humphrey G, Birmingham A, Jepsen K, Farmer S, Tubb HM, Valles T, Tribelhorn CE, Tsai R, Aigner S, Sathe S, Moshiri N, Henson B, Mark AM, Hakim A, Baer NA, Barber T, Belda-Ferre P, Chacon M, Cheung W, Cresini ES, Eisner ER, Lastrella AL, Lawrence ES, Marotz CA, Ngo TT, Ostrander T, Plascencia A, Salido RA, Seaver P, Smoot EW, McDonald D, Neuhard RM, Scioscia AL, Satterlund AM, Simmons EH, Abelman DB, Brenner D, Bruner JC, Buckley A, Ellison M, Gattas J, Gonias SL, Hale M, Hawkins F, Ikeda L, Jhaveri H, Johnson T, et al. 2022. Wastewater sequencing reveals early cryptic SARS-CoV-2 variant transmission. Nature 609:101–108.

13. Smith MF, Holland SC, Lee MB, Hu JC, Pham NC, Sullins RA, Holland LA, Mu T, Thomas AW, Fitch R, Driver EM, Halden RU, Villegas-Gold M, Sanders S, Krauss JL, Nordstrom L, Mulrow M, White M, Murugan V, Lim ES. 2023. Baseline Sequencing Surveillance of Public Clinical Testing, Hospitals, and Community Wastewater Reveals Rapid Emergence of SARS-CoV-2 Omicron Variant of Concern in Arizona, USA. mBio 14:e0310122.

14. Mackenzie JS, Jeggo M. 2019. The One Health Approach-Why Is It So Important? Trop Med Infect Dis 4.

15. O’Brien E, Xagoraraki I. 2019. A water-focused one-health approach for early detection and prevention of viral outbreaks. One Health 7:100094.

16. Xiao K, Zhang L. 2023. Wastewater pathogen surveillance based on One Health approach. Lancet Microbe 4:e297.

17. Boehm AB, Hughes B, Duong D, Chan-Herur V, Buchman A, Wolfe MK, White BJ. 2023. Wastewater concentrations of human influenza, metapneumovirus, parainfluenza, respiratory syncytial virus, rhinovirus, and seasonal coronavirus nucleic-acids during the COVID-19 pandemic: a surveillance study. Lancet Microbe 4:e340–e348.

18. Ahmed W, Bivins A, Stephens M, Metcalfe S, Smith WJM, Sirikanchana K, Kitajima M, Simpson SL. 2023. Occurrence of multiple respiratory viruses in wastewater in Queensland, Australia: Potential for community disease surveillance. Sci Total Environ 864:161023.

19. Toribio-Avedillo D, Gomez-Gomez C, Sala-Comorera L, Rodriguez-Rubio L, Carcereny A, Garcia-Pedemonte D, Pinto RM, Guix S, Galofre B, Bosch A, Merino S, Muniesa M. 2023. Monitoring influenza and respiratory syncytial virus in wastewater. Beyond COVID-19. Sci Total Environ 892:164495.

20. Bibby K, Peccia J. 2013. Identification of viral pathogen diversity in sewage sludge by metagenome analysis. Environ Sci Technol 47:1945–51.

21. Cantalupo PG, Calgua B, Zhao G, Hundesa A, Wier AD, Katz JP, Grabe M, Hendrix RW, Girones R, Wang D, Pipas JM. 2011. Raw sewage harbors diverse viral populations. mBio 2.

22. Martinez-Puchol S, Rusinol M, Fernandez-Cassi X, Timoneda N, Itarte M, Andres C, Anton A, Abril JF, Girones R, Bofill-Mas S. 2020. Characterisation of the sewage virome: comparison of NGS tools and occurrence of significant pathogens. Sci Total Environ 713:136604.

23. Tisza M, Javornik Cregeen S, Avadhanula V, Zhang P, Ayvaz T, Feliz K, Hoffman KL, Clark JR, Terwilliger A, Ross MC, Cormier J, Moreno H, Wang L, Payne K, Henke D, Troisi C, Wu F, Rios J, Deegan J, Hansen B, Balliew J, Gitter A, Zhang K, Li R, Bauer CX, Mena KD, Piedra PA, Petrosino JF, Boerwinkle E, Maresso AW. 2023. Wastewater sequencing reveals community and variant dynamics of the collective human virome. Nat Commun 14:6878.

24. Child HT, Airey G, Maloney DM, Parker A, Wild J, McGinley S, Evens N, Porter J, Templeton K, Paterson S, van Aerle R, Wade MJ, Jeffries AR, Bassano I. 2023. Comparison of metagenomic and targeted methods for sequencing human pathogenic viruses from wastewater. mBio 14:e0146823.

25. Quinn GA, Connolly M, Fenton NE, Hatfill SJ, Hynds P, ÓhAiseadha C, Sikora K, Soon W, Connolly R. 2024. Influence of Seasonality and Public-Health Interventions on the COVID-19 Pandemic in Northern Europe. Journal of Clinical Medicine 13:334.

26. Hamid S, Winn A, Parikh R, Jones JM, McMorrow M, Prill MM, Silk BJ, Scobie HM, Hall AJ. 2023. Seasonality of Respiratory Syncytial Virus - United States, 2017-2023. MMWR Morb Mortal Wkly Rep 72:355–361.

27. CDC. January 11, 2024. The National Respiratory and Enteric Virus Surveillance System (NREVSS). https://www.cdcgov/surveillance/nrevss/indexhtml.

28. Price RHM, Graham C, Ramalingam S. 2019. Association between viral seasonality and meteorological factors. Sci Rep 9:929.

29. Faden H, Wilby M, Hainer ZD, Rush-Wilson K, Ramani R, Lamson D, Boromisa R. 2011. Pediatric adenovirus infection: relationship of clinical spectrum, seasonal distribution, and serotype. Clin Pediatr (Phila) 50:483–7.

30. Lee J, Bilonick RA, Romanowski EG, Kowalski RP. 2018. Seasonal Variation in Human Adenovirus Conjunctivitis: A 30-Year Observational Study. Ophthalmic Epidemiol 25:451–456.

31. Shrestha S, Malla B, Haramoto E. 2023. Estimation of Norovirus infections in Japan: An application of wastewater-based epidemiology for enteric disease assessment. Sci Total Environ 912:169334.

32. Haramoto E, Katayama H, Oguma K, Yamashita H, Tajima A, Nakajima H, Ohgaki S. 2006. Seasonal profiles of human noroviruses and indicator bacteria in a wastewater treatment plant in Tokyo, Japan. Water Sci Technol 54:301–8.

33. Scott MK, Chommanard C, Lu X, Appelgate D, Grenz L, Schneider E, Gerber SI, Erdman DD, Thomas A. 2016. Human Adenovirus Associated with Severe Respiratory Infection, Oregon, USA, 2013-2014. Emerg Infect Dis 22:1044–51.

34. Wang J, Ran L, Zhai M, Jiang C, Xu C. 2023. Prediction of Foodborne Norovirus Outbreaks in Coastal Areas in China in 2008-2018. Foodborne Pathog Dis doi:10.1089/fpd.2023.0037.

35. Abid I, Blanco A, Al-Otaibi N, Guix S, Costafreda MI, Pinto RM, Bosch A. 2023. Dynamic and Seasonal Distribution of Enteric Viruses in Surface and Well Water in Riyadh (Saudi Arabia). Pathogens 12.

36. Winder N, Gohar S, Muthana M. 2022. Norovirus: An Overview of Virology and Preventative Measures. Viruses 14.

37. Bitler EJ, Matthews JE, Dickey BW, Eisenberg JN, Leon JS. 2013. Norovirus outbreaks: a systematic review of commonly implicated transmission routes and vehicles. Epidemiol Infect 141:1563–71.

38. Kitajima M, Sassi HP, Torrey JR. 2018. Pepper mild mottle virus as a water quality indicator. npj Clean Water 1:19.

39. Moriyama M, Hugentobler WJ, Iwasaki A. 2020. Seasonality of Respiratory Viral Infections. Annu Rev Virol 7:83–101.

40. Bowes DA, Driver EM, Kraberger S, Fontenele RS, Holland LA, Wright J, Johnston B, Savic S, Engstrom Newell M, Adhikari S, Kumar R, Goetz H, Binsfeld A, Nessi K, Watkins P, Mahant A, Zevitz J, Deitrick S, Brown P, Dalton R, Garcia C, Inchausti R, Holmes W, Tian XJ, Varsani A, Lim ES, Scotch M, Halden RU. 2023. Leveraging an established neighbourhood-level, open access wastewater monitoring network to address public health priorities: a population-based study. Lancet Microbe 4:e29–e37.

41. Galani A, Aalizadeh R, Kostakis M, Markou A, Alygizakis N, Lytras T, Adamopoulos PG, Peccia J, Thompson DC, Kontou A, Karagiannidis A, Lianidou ES, Avgeris M, Paraskevis D, Tsiodras S, Scorilas A, Vasiliou V, Dimopoulos MA, Thomaidis NS. 2022. SARS-CoV-2 wastewater surveillance data can predict hospitalizations and ICU admissions. Sci Total Environ 804:150151.

42. McMahan CS, Self S, Rennert L, Kalbaugh C, Kriebel D, Graves D, Colby C, Deaver JA, Popat SC, Karanfil T, Freedman DL. 2021. COVID-19 wastewater epidemiology: a model to estimate infected populations. Lancet Planet Health 5:e874–e881.

43. Shah MM, Winn A, Dahl RM, Kniss KL, Silk BJ, Killerby ME. 2022. Seasonality of Common Human Coronaviruses, United States, 2014-2021(1). Emerg Infect Dis 28:1970–1976.

44. DeJonge PM, Adams C, Pray I, Schussman MK, Fahney RB, Shafer M, Antkiewicz DS, Roguet A. 2023. Wastewater Surveillance Data as a Complement to Emergency Department Visit Data for Tracking Incidence of Influenza A and Respiratory Syncytial Virus - Wisconsin, August 2022-March 2023. MMWR Morb Mortal Wkly Rep 72:1005–1009.

45. Wolfe MK, Duong D, Bakker KM, Ammerman M, Mortenson L, Hughes B, Arts P, Lauring AS, Fitzsimmons WJ, Bendall E, Hwang CE, Martin ET, White BJ, Boehm AB, Wigginton KR. 2022. Wastewater-Based Detection of Two Influenza Outbreaks. Environmental Science & Technology Letters 9:687–692.

46. Wolken M, Sun T, McCall C, Schneider R, Caton K, Hundley C, Hopkins L, Ensor K, Domakonda K, Kalvapalle P, Persse D, Williams S, Stadler LB. 2023. Wastewater surveillance of SARS-CoV-2 and influenza in preK-12 schools shows school, community, and citywide infections. Water Res 231:119648.

47. DeCaprio JA, Garcea RL. 2013. A cornucopia of human polyomaviruses. Nat Rev Microbiol 11:264–76.

48. Blackard JT, Davies SM, Laskin BL. 2020. BK polyomavirus diversity-Why viral variation matters. Rev Med Virol 30:e2102.

49. Cortez V, Meliopoulos VA, Karlsson EA, Hargest V, Johnson C, Schultz-Cherry S. 2017. Astrovirus Biology and Pathogenesis. Annu Rev Virol 4:327–348.

50. Esposito S, Principi N. 2020. Norovirus Vaccine: Priorities for Future Research and Development. Front Immunol 11:1383.

51. Simpson S, Kaufmann MC, Glozman V, Chakrabarti A. 2020. Disease X: accelerating the development of medical countermeasures for the next pandemic. Lancet Infect Dis 20:e108–e115.

52. Rader B, Gertz A, Iuliano AD, Gilmer M, Wronski L, Astley CM, Sewalk K, Varrelman TJ, Cohen J, Parikh R, Reese HE, Reed C, Brownstein JS. 2022. Use of At-Home COVID-19 Tests - United States, August 23, 2021-March 12, 2022. MMWR Morb Mortal Wkly Rep 71:489–494.

53. Wang Q, Iketani S, Li Z, Liu L, Guo Y, Huang Y, Bowen AD, Liu M, Wang M, Yu J, Valdez R, Lauring AS, Sheng Z, Wang HH, Gordon A, Liu L, Ho DD. 2023. Alarming antibody evasion properties of rising SARS-CoV-2 BQ and XBB subvariants. Cell 186:279–286 e8.

54. Imai M, Ito M, Kiso M, Yamayoshi S, Uraki R, Fukushi S, Watanabe S, Suzuki T, Maeda K, Sakai-Tagawa Y, Iwatsuki-Horimoto K, Halfmann PJ, Kawaoka Y. 2023. Efficacy of Antiviral Agents against Omicron Subvariants BQ.1.1 and XBB. N Engl J Med 388:89–91.

55. Cole CN. 2001. Polyomaviridae: the viruses and their replication. Fields virology.

56. Pinto M, Dobson S. 2014. BK and JC virus: a review. J Infect 68 Suppl 1:S2–8.

57. Egli A, Infanti L, Dumoulin A, Buser A, Samaridis J, Stebler C, Gosert R, Hirsch HH. 2009. Prevalence of polyomavirus BK and JC infection and replication in 400 healthy blood donors. J Infect Dis 199:837–46.

58. Behzad-Behbahani A, Klapper PE, Vallely PJ, Cleator GM, Khoo SH. 2004. Detection of BK virus and JC virus DNA in urine samples from immunocompromised (HIV-infected) and immunocompetent (HIV-non-infected) patients using polymerase chain reaction and microplate hybridisation. J Clin Virol 29:224–9.

59. Randhawa P, Shapiro R, Vats A. 2005. Quantitation of DNA of polyomaviruses BK and JC in human kidneys. J Infect Dis 192:504–9.

60. Rector A, Bloemen M, Thijssen M, Pussig B, Beuselinck K, Van Ranst M, Wollants E. 2024. Respiratory Viruses in Wastewater Compared with Clinical Samples, Leuven, Belgium. Emerg Infect Dis 30:141–145.

61. Langmead B, Salzberg SL. 2012. Fast gapped-read alignment with Bowtie 2. Nat Methods 9:357–9.

62. Danecek P, Bonfield JK, Liddle J, Marshall J, Ohan V, Pollard MO, Whitwham A, Keane T, McCarthy SA, Davies RM, Li H. 2021. Twelve years of SAMtools and BCFtools. Gigascience 10.

63. Marti-Carreras J, Mineeva-Sangwo O, Topalis D, Snoeck R, Andrei G, Maes P. 2020. BKTyper: Free Online Tool for Polyoma BK Virus VP1 and NCCR Typing. Viruses 12.

64. Haramoto E, Kitajima M, Kishida N, Konno Y, Katayama H, Asami M, Akiba M. 2013. Occurrence of pepper mild mottle virus in drinking water sources in Japan. Appl Environ Microbiol 79:7413–8.

